# Automated detection of bottom-of-sulcus dysplasia on MRI-PET in patients with drug-resistant focal epilepsy

**DOI:** 10.1101/2025.05.08.25326914

**Authors:** Emma Macdonald-Laurs, Aaron E.L. Warren, Remika Mito, Sila Genc, Bonnie Alexander, Sarah Barton, Joseph Yuan-Mou Yang, Peter Francis, Heath R. Pardoe, Graeme Jackson, A. Simon Harvey

## Abstract

**Background and Objectives:** Bottom-of-sulcus dysplasia (BOSD) is a diagnostically challenging subtype of focal cortical dysplasia, 60% being missed on patients’ first MRI. Automated MRI-based detection methods have been developed for focal cortical dysplasia, but not BOSD specifically. Use of FDG-PET alongside MRI is not established in automated methods. We report the development and performance of an automated BOSD detector using combined MRI+PET data.

**Methods:** The training set comprised 54 mostly operated patients with BOSD. The test sets comprised 17 subsequently diagnosed patients with BOSD from the same center, and 12 published patients from a different center. 81% patients across training and test sets had reportedly normal first MRIs and most BOSDs were <1.5cm^3^.

In the training set, 12 features from T1-MRI, FLAIR-MRI and FDG-PET were evaluated using a novel “pseudo-control” normalization approach to determine which features best distinguished dysplastic from normal-appearing cortex. Using the Multi-centre Epilepsy Lesion Detection group’s machine-learning detection method with the addition of FDG-PET, neural network classifiers were then trained and tested on MRI+PET features, MRI-only and PET-only. The proportion of patients whose BOSD was overlapped by the top output cluster, and the top five output clusters, were assessed.

**Results:** Cortical and subcortical hypometabolism on FDG-PET were superior in discriminating dysplastic from normal-appearing cortex compared to MRI features. When the BOSD detector was trained on MRI+PET features, 87% BOSDs were overlapped by one of the top five clusters (69% top cluster) in the training set, 76% in the prospective test set (71% top cluster) and 75% in the published test set (42% top cluster). Cluster overlap was similar when the detector was trained and tested on PET-only features but lower when trained and tested on MRI-only features.

**Conclusion:** Detection of BOSD is possible using established MRI-based automated detection methods, supplemented with FDG-PET features and trained on a BOSD-specific cohort. In clinical practice, an MRI+PET BOSD detector could improve assessment and outcomes in seemingly MRI-negative patients being considered for epilepsy surgery.

## Introduction

Focal cortical dysplasia (FCD) is the most frequent cause of surgically-remediable, drug-resistant, focal epilepsy (DRFE) in young people.^1^ FCD varies in its histological subtypes, imaging characteristics, cortical extent, and clinical manifestations. Whilst some FCDs are easily diagnosed on visual inspection of MRI, due to obvious cortical thickening and grey-white blurring on T1-weighted sequences, or signal hyperintensity on FLAIR and T2-weighted sequences, many are not, even with electroclinical clues to localisation and a 3T epilepsy MRI protocol.^2–4^

The importance of identification, delineation, and complete resection of MRI-visible FCD in patients with DRFE cannot be overstated, given their strong associations with seizure freedom.^5–7^ Failure to detect FCD risks rejection from potentially curative surgery, unnecessary use of intracranial EEG, and erroneous cortical resection, with poor clinical and health economic outcomes.

Sixteen to 43% of patients with DRFE have reportedly normal MRI,^8, 9^ and within this group, bottom-of-sulcus dysplasia (BOSD), the smallest subtype of FCD type 2 (FCDII), is frequently encountered. Approximately 60% of patients with BOSD have their initial MRI reported normal.^2, 10, 11^ BOSD is an important lesion to detect because limited cortical resection results in seizure freedom in 85%, prior intracranial EEG monitoring is usually unnecessary, and delayed diagnosis and treatment risk neurocognitive dysfunction.^2, 12^

Increasingly, automated methods utilizing machine learning are being used to detect FCD on MRI, ranging from large FCD type 1 (FCDI) and mild malformation of cortical development with oligodendroglial hyperplasia and epilepsy (MOGHE), to tiny BOSDs.^13–17^ Such methods include the morphometric analysis program (MAP18)^13, 14^ which uses voxel-based MRI morphometry, the Multi-centre Epilepsy Lesion Detection project (MELD) which uses surface-based MRI morphometry,^15, 16^ and the deepFCD program which uses deep learning on extracted 3D MRI patches.^17^ Most studies report training using FCDs of varying histopathology and size, mostly apparent on MRI, in patients of different ages and with variable epilepsy phenotypes. Although these methods detect some BOSDs,^17^ small FCDs may be misclassified (false negatives) due in part to the underlying clinico-pathological heterogeneity in the training of the automated methods.^13^

Most automated methods for FCD detection train using T1 and FLAIR MRI. While FDG-PET is known to improve visual diagnosis of FCDII,^3^ it is not yet widely utilized in automated detection methods.^18^

We developed an automated detection method for BOSD specifically, using FDG-PET and MRI. We first examined the surface-based MRI and FDG-PET features which best distinguished BOSD from non-dysplastic tissue, then trained the MELD group’s open-source, machine learning, automated detection method^16^ with the addition of FDG-PET feature maps, then finally tested MRI+PET, MRI-only and PET-only detectors on two independent test sets.

## Materials and Methods

### Patients

Patients with BOSD and focal epilepsy managed at the Royal Children’s Hospital (RCH) and the Austin Hospital, Melbourne, Australia between 2008-2024 were included in this study. BOSD was defined as FCD with cortical thickening and blurring of the grey-white junction confined to a single sulcus, maximal at the bottom and tapering to normal overlying gyral crowns, with or without cortical or subcortical signal change.^19^

The training set was derived from a previously described cohort of 85 patients^12^ in whom neuroimaging had been reviewed by four neuroimaging experts to confirm the presence of a BOSD. Patients were excluded if they did not have FDG-PET acquired (n=16), had insufficient quality volumetric FLAIR sequences (n=9), or were younger than two years (n=6) due to poorer Freesurfer^20^ segmentation in this age group.

The test sets comprised 17 children subsequently diagnosed with BOSD at the RCH between 2022-2024 (prospective test set) and a published series of 12 adults diagnosed with BOSD at the Austin Hospital between 2008-2016^21^ (published test set).

Clinical features and imaging details of training and test sets are shown in Table 1.

**Table 1:** Clinical characteristics of training and test sets

|  | **Training set**<br>**(Royal Children's Hospital,**<br>**n=54)** | **Prospective test set**<br>**(Royal Children's Hospital)**<br>**(n=17)** | **Published test set**<br>**(Austin Hospital)<sup>21</sup>**<br>**(n=12)** |
| --- | --- | --- | --- |
| **Sex** | 29 male (54%) | 9 male (53%) | 9 male (75%) |
| **Age at seizure onset (median)** | 6.5 (IQR: 2.4-8.6) years | 8.2 (IQR: 4.1-12.2) years | 5.5 (IQR: 3-9.3) years |
| **Age at MRI scan (median)** | 10.8 (IQR: 7.4-13.4) years | 13.1 (IQR: 10-14.1) years | 28.6 (IQR: 23.2-36.6) years |
| **BOSD reported on first MRI scan** | 10 (19%) | 4 (24%) | unknown |
| **BOSD location** |  |  |  |
| **Frontal** | 28 (52%) | 13 (78%) | 9 (75%) |
| **Parietal** | 13 (24%) | 1 (5.6%) | 3 (25%) |
| **Insula** | 8 (15%) | 3 (17%) | 0 |
| **Temporal** | 5 (9%) | 0 | 0 |
| **Period neuroimaging performed** | 2012-2022 | 2023-2024 | 2008-2016 |
| **Epilepsy surgery** | 41 (76%) | 8 (47%)<sup>a</sup> | 12 (100%)<sup>b</sup> |
| **Histopathology subtypes** |  |  |  |
| **FCDIIA** | 18 (44%) | 4 (50%) | 1 (8%) |
| **FCDIIB** | 21 (51%) | 2 (25%) | 11 (92%) |
| **no pathology** | 2 (5%) | 2 (25%) |  |

<sup>a</sup>four other patients are awaiting surgery or completing pre-surgical work-up, <sup>b</sup>one patient had thermocoagulation following stereo-EEG implantation and one patient MRI-guided laser interstitial thermal therapy

### MRI and FDG-PET acquisition

MRIs were acquired at 3T using an epilepsy protocol,^2^ including T1-weighted volumetric (0.8-0.9 mm isotropic voxels), and FLAIR volumetric (0.5×0.5×0.9 mm^3^) sequences. FDG-PET scans were acquired with a slice thickness of 2mm and FDG uptake time of 30 minutes.

MRIs in the training set were mostly (69%) acquired on a Siemens mMR Biograph with simultaneous FDG-PET acquisition. The remainder of the training set patients had MRI and FDG-PET acquired separately (Supplementary Table 1). The prospective test set patients all had MRIs acquired on the Siemens mMR Biograph concurrently with FDG-PET. MRI and FDG-PET were acquired across several scanners in the published test set (Supplementary Table 1). FLAIR sequences in the published test set patients were either not acquired volumetrically or Freesurfer (*recon-all*) failed, meaning only T1 and FDG-PET data were used in this test set.

### BOSD segmentation and MRI pre-processing

For all patients, the dysplastic-appearing cortex of each BOSD was manually segmented by one author (EML) using MRview within MRtrix3 software using T1-weighted orthogonal slices with FLAIR overlay.^22, 23^ Smoothing with a 2mm full width at half-maximum (FWHM) Gaussian kernel was applied to each drawing to minimize minor drawing inaccuracies.

FreeSurfer (version 7.1.1)^20^ was used to generate surface reconstructions of each patient’s brain using the *recon-all* function, using FLAIR sequences, if available.^20^ FLAIR-MRI and FDG-PET images were aligned to the T1-MRI and then projected to the cortical surface. A nonlinear warp was calculated from each patient’s native cortical surface to a symmetric template surface (*fsaverage_sym*) using FreeSurfer’s *Xhemi* procedure.^24^

### Quantitative analysis of BOSD surface features

Using the training set, surface-based features of BOSD on T1-MRI, FLAIR-MRI, and FDG-PET were analyzed to firstly determine which features were most useful in distinguishing BOSD from normal-appearing brain.

FreeSurfer was used to calculate 12 measures of cerebral morphology, T1 and FLAIR signal, and metabolism, similar to those described by the MELD group^16, 25^ but with addition of PET features (Supplementary Table 2). For each measure, feature maps across the cortical surface were generated, spatially smoothed, and normalized using within-subject *z*-scoring to adjust for inter-individual differences in the global mean and standard deviation.

Given no healthy controls were available, a novel approach was used to define a patient-specific group of “pseudo-controls” derived from the training set. For each patient, we generated a group of pseudo-controls to compare their BOSD with “normal” appearing cortex. Each patient’s pseudo-control group had to meet two criteria: (i) they did not have a BOSD in the same hemisphere as the patient,^23, 26^ and (ii) they did not have a BOSD in the contralateral parcel of the patient’s BOSD as defined by the Desikan-Killiany cortical parcellation.^27^ The median number of pseudo-controls for each patient was 26 (IQR: 24-27).

To assess the degree to which each imaging feature distinguished dysplastic from normal-appearing cortex, we compared each patient’s BOSD to the same cortical location defined either (i) ipsilaterally, using the average across the matched group of pseudo-controls, across vertices within the patient’s lesion drawing; or (ii) contralaterally, using a mirror cortical location in the patient’s non-BOSD hemisphere; this mirroring was achieved using FreeSurfer’s *Xhemi* registration.^24^ The one-sample Kolmogorov-Smirnov test of normality was used to determine whether data were normally distributed. A paired *t*-test was used for normally distributed data and a Wilcoxon signed rank test for non-normally distributed data (local cortical deformation and FLAIR white matter signal). Significance was assessed at *p*<0.05, Bonferroni-corrected for 12 features (α_original_ = 0.05 α_corrected_ = 0.004). These comparisons were performed using R Studio V1.4.1106.

### Training the MELD group’s FCD detection method with a BOSD set and addition of FDG-PET

We next sought to determine if the MELD group’s automated FCD detection code could be trained to detect BOSD, given their small size, and whether addition of FDG-PET features would improve detection. For this analysis, using the training set, each patient’s within-subject *z*-scored maps were further normalized by the mean and standard deviation across their matched group of pseudo-controls (i.e., between-subject *z-*scoring), similar to the original MELD paper^16^ (Figure 2).

**Figure 1.**
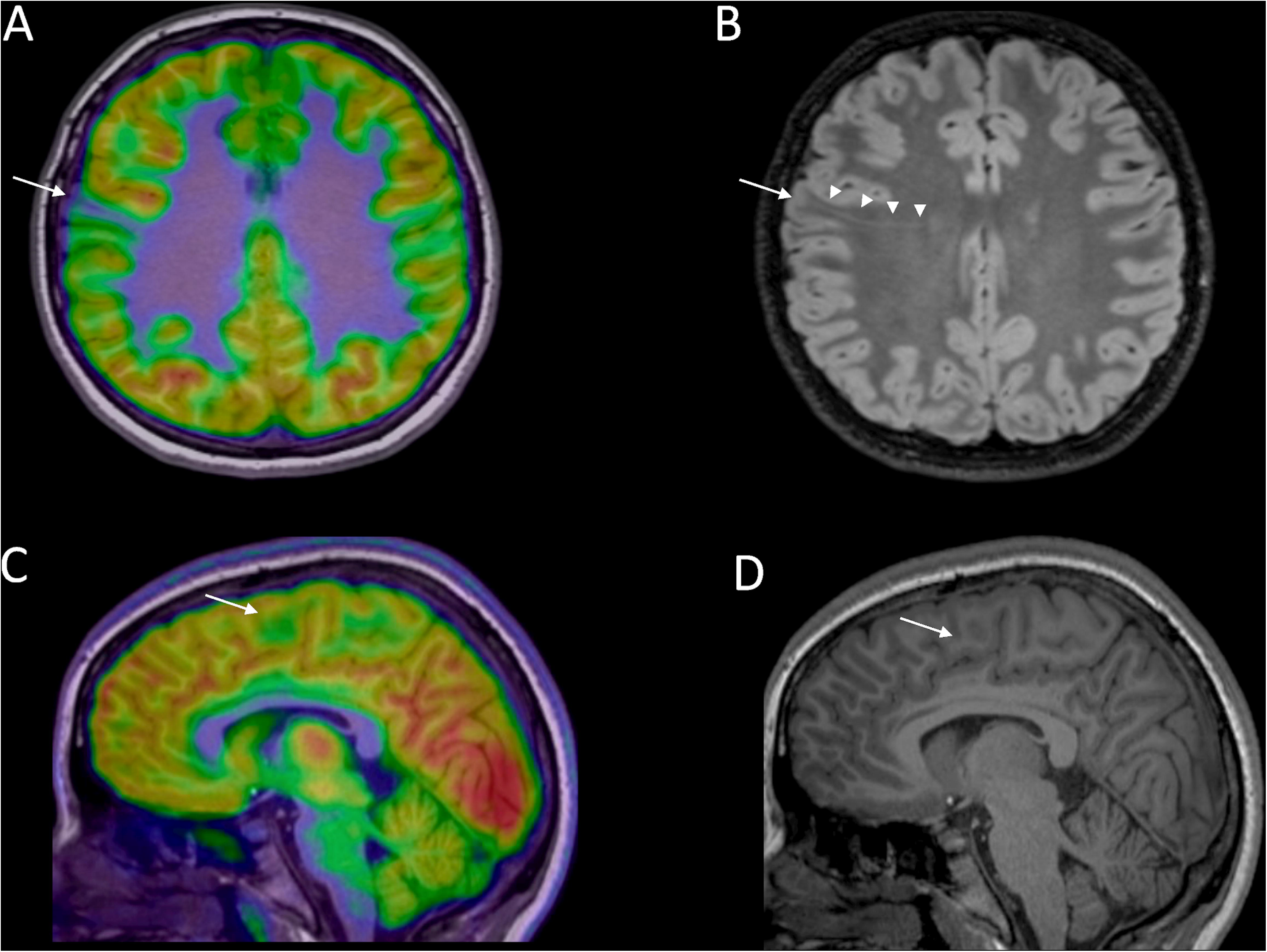
Example images of two patients included in the training set with BOSDs that were not reported on their first MRI but were later identified visually following FDG-PET co-registered with MRI. **A** and **B** show a BOSD in the right subcentral gyrus detected following two “negative” epilepsy protocol MRIs in an school-aged male following review of his FDG-PET scan. A focal region of cortical hypometabolism is seen on FDG-PET co-registered with T1-weighted MRI in axial view (**A**). On axial FLAIR MRI (**B**), grey-white blurring of the bottom of a shallow sulcus is seen (arrow) as well as a feint transmantle band (arrowheads). This BOSD was resected with FCDIIA histopathology identified. He remains seizure free off medication at 3.2 years follow-up. **C** and **D** show a BOSD in the medial surface of the superior frontal gyrus which was missed on several epilepsy protocol MRIs over 10 years following seizure onset in an adolescent female. FDG-PET co-registered with T1 MRI in sagittal view (**C**) shows a subtle focal region of cortical and subcortical hypometabolism (arrow). On sagittal T1 MRI (**D**) subtle cortical thickening and blurring is evident at the bottom of the sulcus (arrow). This BOSD was resected with FCDIIB histopathology identified. She remains seizure free off medication at two years follow-up.

**Figure 2:**
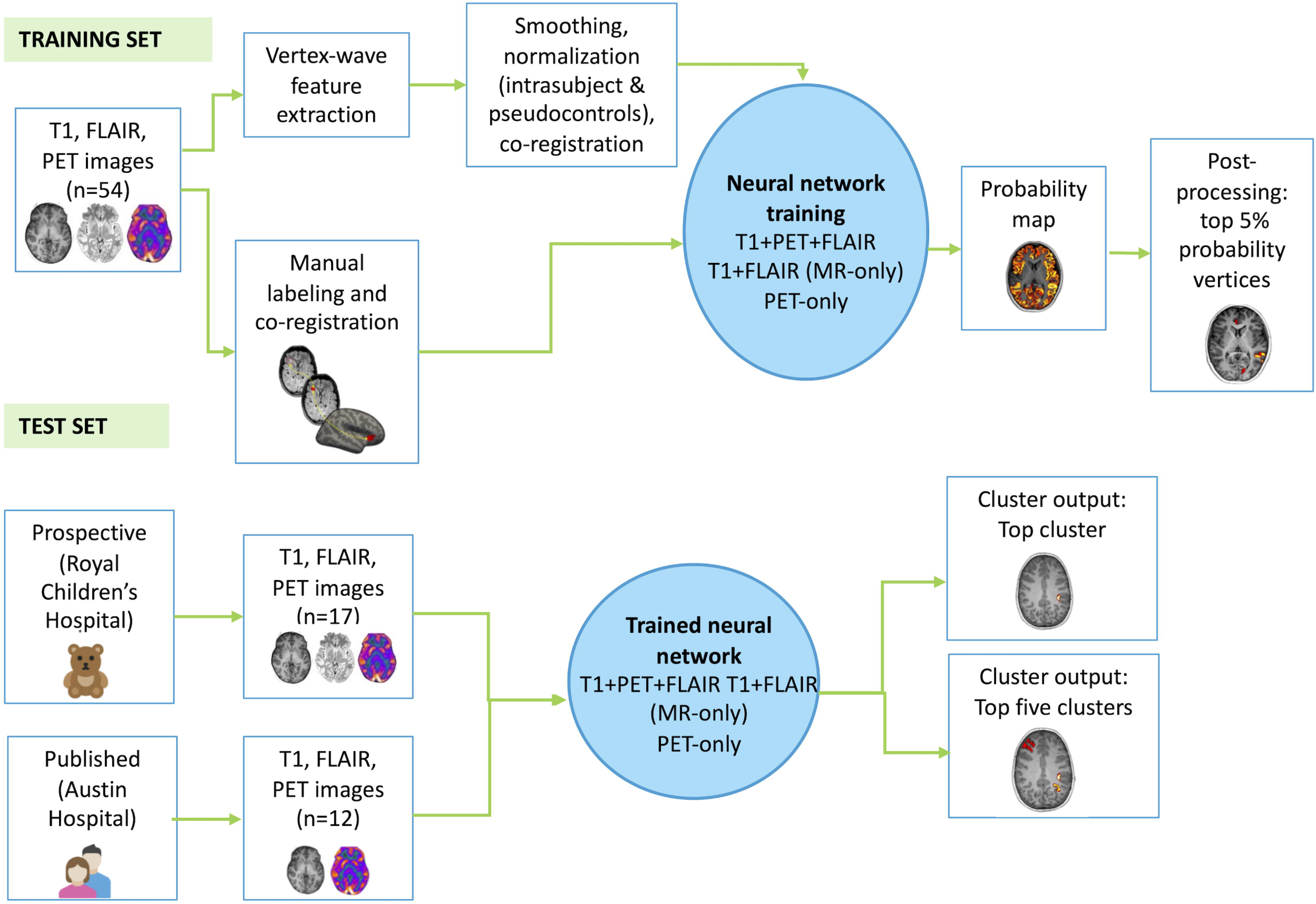
MRI+PET BOSD detector pipeline. Using the training set (n=54), surface-based morphological features from T1-MRI, FLAIR-MRI and FDG-PET were extracted and BOSD were manually segmented and then registered to common template space. Both intra and intersubject normalization steps were performed, the latter using a novel pseudo-control approach. The neural network classifier was trained to identify lesional vertices and was evaluated using a leave-one-out cross validation technique. The trained neural network was tested on a similarly aged prospective pediatric BOSD cohort (n=17) and an historical adult BOSD cohort (n=12). Post processing steps produced visual lesion probability maps (top cluster and top five clusters) for visual review.

A single hidden layer neural network classifier^16^ was trained using the Neural Network Toolbox in MATLAB version 2020b, following steps outlined in the open-access code provided by the MELD authors (https://github.com/kwagstyl/FCDdetection/).^16^ The detector was trained using a supervised learning approach, using the pseudo-control-normalized MRI and FDG-PET feature maps from each patient as input features trained to predict the binary BOSD segmentation maps as output (1=BOSD and 0=not BOSD). The number of network nodes was determined by principal component analysis of all input feature maps combined, with the node number set to the number of components explaining >99% of the variance.^16^

All 12 input features from T1-MRI, FLAIR-MRI and FDG-PET were included in the main analysis. Additionally, separate classifiers were trained using MRI-only (T1 + FLAIR, 10 features), and PET-only (2 features), to assess the discriminatory value of specific imaging modalities. Prior to evaluation on the independent held-out dataset, the performance of each detector was internally assessed using the training dataset using a leave-one-out cross-validation approach.^28^

The output of the neural network classifier was a probability map where values closer to 0 were more likely to be non-BOSD cortex and values closer to 1 were more likely to be BOSD cortex. For each subject, the probability map was thresholded so that only the top 5% of vertices remained, producing a set of candidate clusters.^16^ Small clusters of fewer than 200 vertices were excluded. We also explored consistency of results using other thresholds (see Results).

For each detector, accuracy was defined as the percentage of tested subjects whose BOSD segmentation overlapped (i) the “top cluster”, that being the cluster with the highest mean probability value, and (ii) one of the top five clusters. The mean number of clusters overlapping the hemisphere contralateral to the lesion is reported as a surrogate measure of specificity.

Mann-Whitney U testing was performed to compare clinical features between subjects whose BOSDs were either detected or not detected.

### Testing the MRI-PET BOSD detector on independent test sets

Finally, we tested the BOSD detectors developed above on the independent test sets. For the published test set, given no FLAIR images were available the T1-MRI-only, and T1-MRI+PET detectors were used. Accuracy was defined in the same manner as described for the training set.

### Ethics approval

The study was approved by Human Research and Ethics Committee at the Royal Children’s Hospital (HREC 36328) and Austin Hospital (2013/05123).

### Data availability

Anonymized data not published within this article will be made available upon reasonable request.

## Results

BOSD locations in the training and test sets were similar (Table 1). Median BOSD volume in the training set was 1.25 (IQR: 0.47-2.3) with 25 BOSDs being less than 1cm^3^ in size. The median BOSD volume of the prospective test set was 0.73 (IQR: 0.30-1.68) cm^3^ and 1.83 (IQR: 1.24-3.81) cm^3^ for the published test set. Apart from age at scan and lesion volume for the published test set, patient demographics were similar in the training and test sets.

### Surface features differentiating BOSD from normal brain

When compared to both the ipsilateral location in pseudo-controls and the contralateral hemisphere mirror location within each patient in the training set, the features showing the greatest difference between the BOSD and comparator were, in descending order of statistical significance: decreased cortical metabolism, decreased subcortical metabolism, increased subcortical FLAIR signal, decreased T1 grey-white matter contrast, and increased cortical T1 signal (all *p*<0.0001, Bonferroni corrected) (Figure 3). Lower ranked but significant differences in z-score (*p*<0.05, Bonferroni corrected) were also found for sulcal curvature, sulcal depth, cortical thickness, and subcortical FLAIR signal. Local cortical deformation was significantly different when compared to the contralateral hemisphere, but not to pseudo-controls. Local gyrification index and T1 subcortical white matter signal were not significantly different on either comparison (Figure 3).

**Figure 3.**
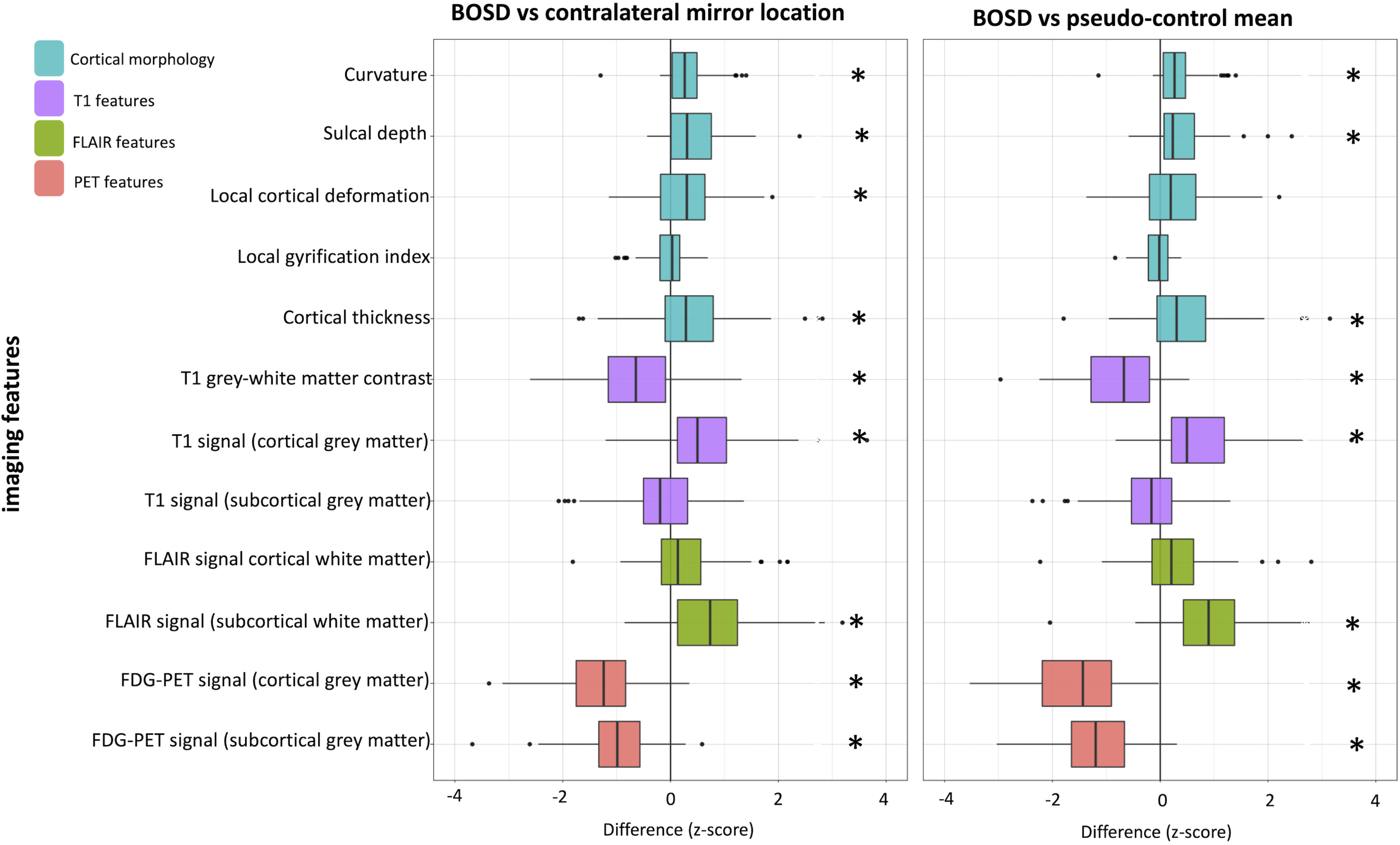
Boxplots of the mean *z*-scores of the 12 measured features compared to the patients’ contralateral hemispheres (left) or a group of pseudo-controls (right) in the training set (n=54). Boxes indicate the data between the 25^th^ and 75^th^ percentile. Black dots represent outliers. Stars denote measures which were significant at *p*<0.05 (Bonferroni corrected). The five features showing the greatest difference between the BOSD, and comparator were, in descending order: decreased cortical metabolism, decreased subcortical metabolism, increased subcortical FLAIR signal, decreased T1 grey-white matter contrast, and increased T1 signal.

### Assessment of the MELD group’s automated FCD detection method trained with BOSD patients and incorporation of FDG-PET

Examples of cluster probability maps are shown in Figures 4 and 5. When the detector was trained on MRI+PET features, one of the top five clusters overlapped the BOSD in 47 (87%) patients in the training set, with the “top cluster” overlapping in 37 (69%) (Figure 6). When the detector was trained on MRI-only, one of the top five clusters overlapped the BOSD in 61% patients (top cluster: 59%) and when it was trained on PET-only, one of the top five clusters overlapped in 83% patients (top cluster:74%).

**Figure 4.**
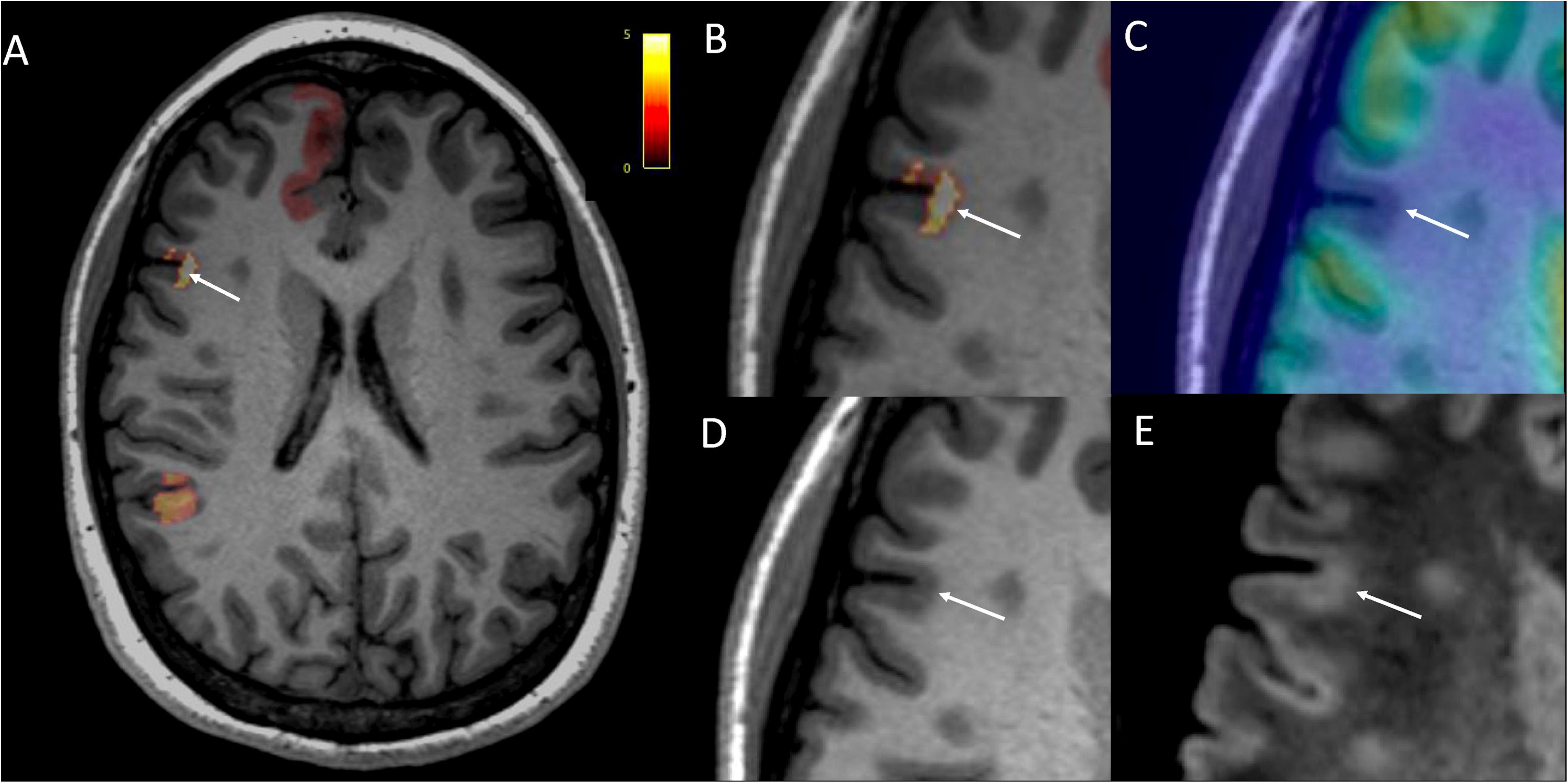
Example of a cluster probability map output of a detector trained on all 12 features in an adolescent female in the test set with a pathologically proven FCDIIB BOSD in the right inferior frontal gyrus. The cluster probability map shows 3 of the top 5 clusters on the patient’s T1-weighted axial MRI (**A, B** zoomed) with the top cluster highlighted (arrow). The patient’s FDG-PET scan co-registered with T1-weighted MRI demonstrates marked cortical and mild subcortical hypometabolism at the bottom of the sulcus, extending superiorly to involve the overlying gyral crown (**C**). On a zoomed view, cortical thickening and slight blurring of the grey-white matter junction is evident on the axial T1-weighted MRI (**D**) and increased cortical signal intensity is seen on the axial FLAIR MRI (**E**). No imaging abnormality was confirmed visually in the other two clusters displayed.

**Figure 5.**
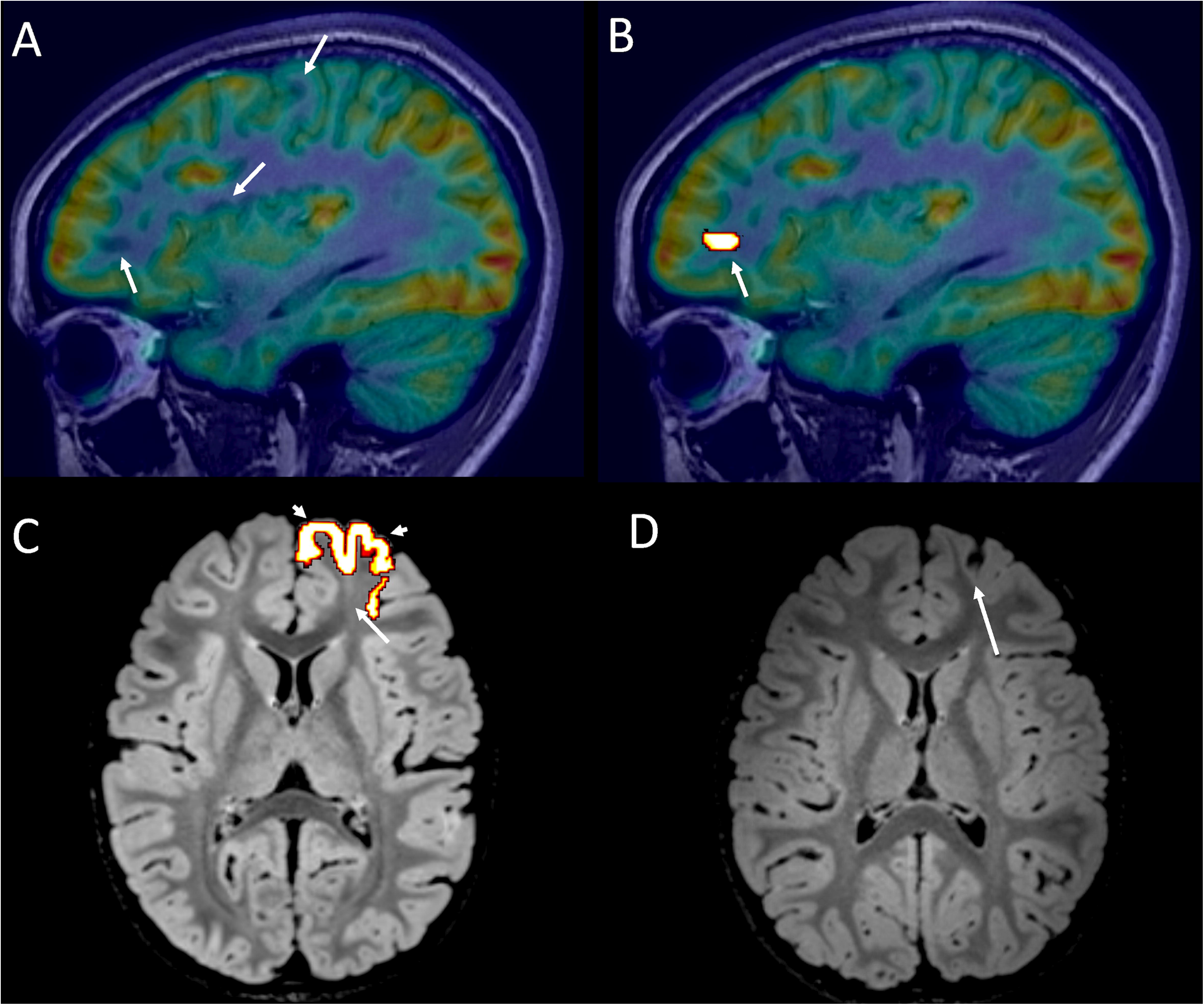
Example of the how cluster outputs are interpreted for two patients in the prospective test set. The top row shows an adolescent female with MRI-negative drug-resistant frontal lobe epilepsy with non-localizing seizure semiology and EEG findings had multiple regions of FDG-PET hypometabolism (arrows) appreciated on visual review of sagittal FDG-PET co-registered to T1 MRI, including in the right inferior frontal gyrus, right superior circular sulcus and right superior frontal gyrus (**A)**. The top cluster output of the PET+MRI BOSD detector overlapped one of the hypometabolic regions, in the right inferior frontal sulcus (arrow), but did not highlight the other regions (**B**). On closer visual inspection, subtle T1 pallor and blurring were appreciated at the bottom of this sulcus. Given the subtle MRI findings, stereo-EEG was performed at an adult center six months later, confirming this right inferior frontal sulcus region to be the epileptogenic zone. Radiofrequency thermocoagulation of the presumed BOSD resulted in sustained seizure freedom. The bottom row shows the top cluster output of a preschool-aged male with multiple hourly gelastic seizures is shown on axial FLAIR MRI (**C**). This cluster overlaps a BOSD with a feint transmantle band (arrow) as well as subtle cortical thickening and grey-white blurring in the left frontal pole. The cluster also overlaps relatively normal appearing gyral crown (arrowheads). Given MRI abnormalities were localized at the bottom of the sulcus, the patient underwent a targeted, sulcus only resection (**D**) which revealed FCDIIB histopathology. He remains seizure free 12 months later, off medication. This example demonstrates that a cluster output may be larger (or potentially smaller) than the BOSD or FCD and should be used only to localize a lesion, rather than determine its possible extent or resection margin.

**Figure 6.**
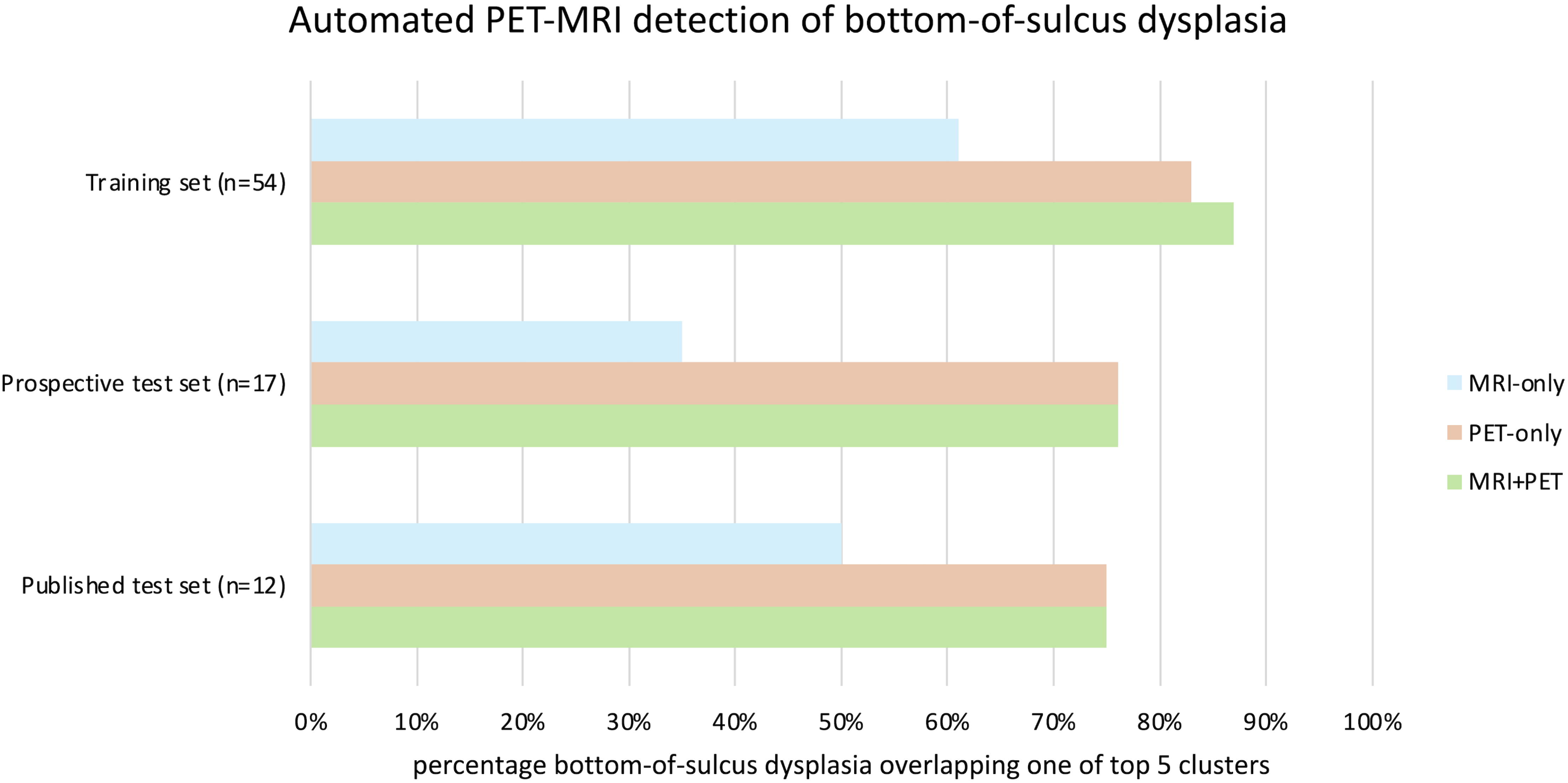
Histogram demonstrating the percentage of BOSD which were overlapped by one of the top five clusters for the detector trained on MRI+PET (green), PET-only (apricot), MRI-only (sky blue) for the RCH training set (n=54), the RCH test set (n=17), and Austin test set (n=12).

The mean number of clusters overlapping the contralateral hemisphere was 1.62 (SD: 0.6) for the MRI+PET detector, 1.66 (SD: 0.48) for the PET-only detector and 2.1 (SD: 0.62) for the MRI-only detector.

Patients whose BOSD was not overlapped with one of the top five clusters on the MRI+PET detector had smaller BOSD volume (Mann-Whitney U, *z*=2.04, *p*=0.04) compared to patients whose BOSDs overlapped. Younger age at scan, being reported as MRI-negative, and FCDIIA histopathology subtype were not associated with lower detection rate. To ensure BOSDs were not being missed by the detector due to thresholding of the probability map, we performed a sensitivity analysis with thresholds excluding clusters less than 50, 100, 150, 200, 250, 300, 350 and 400 vertices. No difference in the rate of cluster overlap was found between different thresholds.

### Performance of the automated method for BOSD detection in independent test sets

For the MRI+PET detector, BOSDs in the prospective test set were overlapped by one of the top five clusters in 76% patients (top cluster: 71%) (Figure 6, Supplementary Table 1). In the published test set, BOSDs were overlapped by one of the top five clusters in 75% patients (top cluster: 42%).

Overlap was slightly less for the PET-only detector, with 76% patient’s BOSDs overlapped in the prospective test set for one of the top five clusters (top cluster: 65%) and 75% in the published test set (top cluster: 42%).

Overlap for the MRI-only detector was lower with 35% patient’s BOSD overlapped by one of the top five clusters in the prospective test set (top cluster: 24%) and 50% overlapped in the published test set (top cluster: 42%).

## Discussion

We showed using previously described, surface-based methods^16^ that automated FCD detection is possible in patients with tiny BOSDs when the detector is trained on a BOSD cohort and FDG-PET features supplement MRI-based features. Most BOSDs were less than 1.5cm^3^ in volume and 80% were not diagnosed on their first MRI, confirming the small and subtle nature of these lesions in the training and test sets and the potential clinical utility of such a detector.

We showed that decreased cortical and subcortical metabolism on FDG-PET was a stronger discriminator of dysplastic cortex than 10 surface-based MRI features. Addition of FDG-PET to the MELD group’s surface-based, FCD detection method improved detection of BOSDs in the training set beyond MRI features alone, increasing the detection rate from 61% to 87% when the top five clusters were reviewed. The detection rate of the MRI+PET detector in the prospective pediatric test set was similar, with 76% patient’s BOSDs overlapped by one of the top five clusters (top cluster: 71%). Detection rate was lower, but still reasonable, in the published adult test set, potentially due to different age at scan acquisition, non-harmonization of imaging data, lack of 3D FLAIR sequences for Freesurfer cortical reconstruction and automated detection steps, and possibly more extensive hypometabolism in an adult cohort. This cohort was chosen as it was one of few published BOSD series.

### Supplementing MRI-based FCD detection with hypometabolism from FDG-PET

The finding that cortical hypometabolism was the most significant feature discriminating BOSDs from their normal comparator is unsurprising given FDG-PET is highly sensitive for visually detecting FCDII, particularly when co-registered or co-acquired with MRI.^3, 29, 30^ In an early series of MRI-negative FCD patients, Chassoux and colleagues reported that focal cortical hypometabolism aided visual FCD detection, including in eight patients in whom hypometabolism was limited to the depth of a single sulcus.^3^ Since then, other FCD series, including BOSD series, have corroborated this observation.^2, 21, 23, 29^

Our MRI-PET and PET-only detectors had sensitivities that were comparable to other MRI-only detectors in larger lesion FCD cohorts,^15–18^ particularly when the top five clusters were considered. The PET-only detector outperformed MRI in the BOSD training set, this potentially being due to MRI abnormalities in BOSDs being of very small volume and difficult to distinguish (even visually) from normal cortex, compared to larger volume cortical hypometabolism (Figure 1). Despite BOSDs almost universally having reduced cortical metabolism at the sulcal depth,^12^ FDG-PET scans are sometimes reported as normal, even with MRI coregistration (Figure 4). Reasons for this include small BOSD size and hypometabolism being confined to the depth of the sulcus with contiguous normal signal occurring over the gyral crown, this hindering visual detection. Automated approaches to FDG-PET analysis overcome these limitations and so may add value compared to visual analysis alone.

Recently, other groups also reported that addition of FDG-PET to automated FCD detection methods is usually advantageous. Similar to our findings, Tan et al. showed in 28 patients with mixed FCDI, FCDIIA and FCDIIB histopathology that the sensitivity of lesion detection improved when FDG-PET was added to their detector, from 82% to 93%; however their FDG-PET-only detector had a lower sensitivity of 61%.^31^ Interestingly, for the 11 patients with FCDIIA histopathology, only FDG-PET features discriminated lesional from non-lesional cortex.^31^ This is of particular importance in patients with BOSD, as almost half have FCDIIA histopathology, especially in “MRI-negative” group. More recently, using a novel volume-based 3D convolutional neural network method, Zheng et al. showed that addition of FDG-PET to MRI increased FCD detection in a mixed FCDI and FCDII cohort, in both training and test sets, with high sensitivities.^18^ In contrast, Mo et al. reported that addition of FDG-PET data to a similar surface-based method in 74 patients did not improve FCD detection compared to MRI alone, this potentially relating to the use of hippocampal sclerosis as a control group in this study.^32^

Cortical hypometabolism on FDG-PET relates to both the pathological lesion and to local and remote network dysfunction. For example in FCDIIB lesions, cortical hypometabolism occurs locally due to abnormal uptake of FDG by dysmorphic neurons and balloon cells, while perilesional and remote hypometabolism may be impacted by seizure duration, frequency, and severity.^3, 23, 33^ Regions of cortical hypometabolism in patients with DRFE have been shown to overlap with markers of the epileptogenic zone including spiking, high frequency oscillations and ictal onset on intracranial EEG, and regions of hyperperfusion on ictal SPECT.^34, 35^ We reported in BOSD specifically that maximal cortical hypometabolism overlaps maximal epileptogenicity, cortical thickness, dysmorphic neuron density and *MTOR* somatic variant allele frequency.^23^ Although the extent of cortical hypometabolism is often larger than the epileptogenic zone,^29^ several studies have shown a gradient of improving cortical metabolism, along with decreasing epileptogenicity, as the distance from the FCDII nidus increases. These observations support the idea that measurement of regions of maximal hypometabolism in an FCD detector may aid in lesion detection, even if, and potentially because, it may be more extensive than the pathological lesion and sensitive to factors in addition to pathology.^23, 34, 35^

The finding that subcortical hypometabolism was also highly discriminative is intriguing. This may reflect decreased FDG uptake in abnormal oligodendroglial and astroglial cells, these cells potentially having a role in “infra-slow” activity measured in the epileptogenic zone on intracranial EEG.^36–38^ Alternatively, subcortical hypometabolism may occur as a result of altered white matter microstructure resulting from frequent seizures.^39–41^ Another possibility is that the subcortical hypometabolism reflects signal spread away from, or partial voluming with, the BOSD itself.

### Patient and dysplasia specific training of FCD detectors

Significant progress in automated FCD detection has occurred in recent years, with several multicenter studies reporting sensitivities between 59-83% on independent test data sets.^13, 15, 17, 42^ To date, most FCD detectors have been trained on heterogenous clinical cohorts with MRI-positive and MRI-negative lesions of variable size and histopathology, affording some advantages and disadvantages in test situations.^13, 15, 17, 42^ Our training and test sets were restricted to patients with small BOSDs, most with proven FCDII histopathology and a common underlying mTORopathy genetic basis. Additionally, nearly all patients presented with the electroclinical features characteristic of BOSD, i.e. childhood-onset unifocal seizures, normal intellect, and focal epileptiform discharges.^12^ Generalized seizures including epileptic spasms, generalized EEG abnormalities, and intellectual disability are common features of some childhood-onset, FCD-associated epilepsies, but not BOSD. In addition to the incorporation of FDG-PET features, training and testing in homogeneous clinico-pathological patient groups likely contributed to the good performance of our MRI+PET BOSD detector, despite small lesion size.

Given the rapid improvements in model building and architecture, there is increasing emphasis on data selection and curation to improve model outputs.^43^ The approach of training FCD detectors on patient and pathology specific cohorts could be valuable in other cohorts, for example in infants with epileptic spasms who have a higher probability of having larger but still subtle FCDI or MOGHE lesions. Although not formally tested, our in-house experience is that our MRI+PET BOSD detector performs less well in this patient group, again suggesting the need for development of radiologically, histopathologically, or phenotypically targeted detectors.

### Clinical application of automated FCD detection

Increasingly, clinicians will use outputs from automated methods in their review of MRI for subtle FCD, in conjunction with clinical, EEG and functional imaging data. From a clinical application perspective, MRI+PET BOSD detector outputs should direct the radiologist to review MRI images more closely for visual characteristics of BOSD and reformat images for closer scrutiny if necessary. Additionally, detector outputs might corroborate or de-emphasize cortical areas that were suspicious for a BOSD on visual inspection, by providing independent, objective analysis. Presently, BOSD should only be diagnosed when characteristic MRI features are confirmed on visual review, and patients should proceed to surgery only if the lesion and its location are consistent with clinical and EEG data.

Output of an MRI+PET BOSD detector can result in an “MRI-negative” patient becoming “+I-positive”, this having clinical implications including: consideration as a surgical candidate, avoidance of intracranial EEG monitoring,^2^ being able to perform targeted resection or ablation,^23^ or contributing to intracranial EEG planning when necessary.^44^ At our center, increasing use of neuroimaging post-processing methods and the MRI+PET BOSD detector have resulted in both an increased rate of BOSD diagnosis, decreased duration from presentation to diagnosis,^12^ and decreased utilization of intracranial EEG monitoring.

Automated methods have their limitations in clinical practice. Limitations may be related to the model itself, including its sensitivity, specificity or data transparency. Clinical application of FCD detectors may be impeded by technical factors, for example, lack of programming skills or the need to perform harmonization across different MRI scanners. Most importantly FCD detectors, particularly detectors trained on specific lesions like ours, should be applied to the correct patient groups. The output of FCD detectors should not replace the skills of neuroradiologists and epileptologists, but instead be used to complement and inform hypotheses and formulations.

### Study Limitations

Availability of control FDG-PET data is a challenge in children, due to differences in myelination and FDG uptake across development, radiation exposure, and sometimes need for anesthesia. Our novel method of forming a group of patient-specific, pseudo-controls attempted to overcome the issue of not having typically developing controls. Other studies used patients with medial temporal lobe epilepsy as FDG-PET controls,^32^ this potentially introducing bias.^33^ Non-epilepsy patients, typically oncology patients, are problematic FDG-PET controls due to briefer brain acquisition, effects of steroids and chemotherapies, and potential cerebral malignancies.

No MRI harmonization correction steps were performed between the various MRI and FDG-PET scanners used, due to the small cohort.^25, 45^ Lack of harmonization may have reduced detection, particularly for the published test set. However, addition of FDG-PET to an FCD detector may reduce the need for harmonization steps relative to use of MRI features only, especially when detectors are to be used in expanded clinical cohorts.

Our study is limited by small cohort size, relative to other studies which included large cohorts from multiple centers, potentially resulting in class imbalance. However, our cohort is strengthened by its clinical, imaging, pathological, and genetic homogeneity, and by the high proportion of lesions having proven pathology and localization following surgery and subsequent seizure freedom.

Our future work will focus on re-training the MRI+PET BOSD detector on a greater number of more recent, more subtle, and all pathologically proven BOSD, incorporating explainable machine learning approaches to better understand the contribution of PET at an individual patient level, and finally prospective evaluation of the detector in non-harmonized multicentre cohorts.

## Conclusion

We showed that automated detection of small and often overlooked BOSD using publicly available, surface-based methods developed from a heterogeneous FCD cohort is possible, with addition of FDG-PET features and training on a BOSD cohort. ^16, 25^ The clinical utility of such an MRI+PET BOSD detector needs to be tested in prospective cohorts of patients with DRFE whose electroclinical features are suggestive of BOSD but MRI is either negative, inconclusive or subtle. The potential benefits in epilepsy surgery are significant, including improving patient selection, strengthening seizure localisation, limiting surgical resection, and leading to better clinical and health economic outcomes.

## Acknowledgements

We thank and acknowledge the Multi-centre Epilepsy Lesion Detection (MELD) project team members, particularly the MELD team leaders Dr Sophie Adler & Dr Konrad Wagstyl, who developed the methods used in this work and provided advice.

We thank our colleagues Michael Kean, Duncan Veysey, Trish Cahill, Kathryn Santamaria, Dr Cristina Mignone, Dr John Fitzgerald, A/Professor Simone Mandelstam, Dr David Vaughn, Dr Mira Semmelroch and Dr Fiona Permezel for their contributions to clinical image acquisition and processing.

This research was conducted within the Developmental Imaging research group, Murdoch Children’s Research Institute, Royal Children’s Hospital, Melbourne Victoria. It was supported by the Murdoch Children’s Research Institute, Royal Children’s Hospital, The University of Melbourne Department of Paediatrics and the Victorian Government’s Operational Infrastructure Support Program. The authors acknowledge the facilities and scientific and technical assistance of the National Imaging Facility, a National Collaborative Research Infrastructure Strategy (NCRIS) capability.

## References

1. Widjaja E, Jain P, Demoe L, Guttmann A, Tomlinson G, Sander B. Seizure outcome of pediatric epilepsy surgery. Neurology. 2020;94:311–321.

2. Macdonald-Laurs E, Maixner WJ, Bailey CA, et al. One-stage, limited-resection epilepsy surgery for bottom-of-sulcus dysplasia. Neurology. 2021;97:e178–e190.

3. Chassoux F, Rodrigo S, Semah F, et al. FDG-PET improves surgical outcome in negative MRI Taylor-type focal cortical dysplasias. Neurology. 2010;75:2168–2175.

4. Chassoux F, Landré E, Mellerio C, et al. Type II focal cortical dysplasia: Electroclinical phenotype and surgical outcome related to imaging. Epilepsia. 2012;53:349–358.

5. Krsek P, Maton B, Jayakar P, et al. Incomplete resection of focal cortical dysplasia is the main predictor of poor postsurgical outcome. Neurology. 2009;72:217–223.

6. McIntosh AM, Averill CA, Kalnins RM, et al. Longlterm seizure outcome and risk factors for recurrence after extratemporal epilepsy surgery. Epilepsia. 2012;53:970–978.

7. Englot DJ, Wang DD, Rolston JD, Shih TT, Chang EF. Rates and predictors of long-term seizure freedom after frontal lobe epilepsy surgery: a systematic review and meta-analysis: Clinical article. Journal of Neurosurgery JNS. 2012;116:1042–1048.

8. Bien CG, Szinay M, Wagner J, Clusmann H, Becker AJ, Urbach H. Characteristics and surgical outcomes of patients with refractory magnetic resonance imaging-negative epilepsies. Arch Neurol. 2009;66:1491–1499.

9. McGonigal A, Bartolomei F, Régis J, et al. Stereoelectroencephalography in presurgical assessment of MRI-negative epilepsy. Brain. 2007;130:3169–3183.

10. Snyder K, Whitehead EP, Theodore WH, Zaghloul KA, Inati SJ, Inati SK. Distinguishing type II focal cortical dysplasias from normal cortex: A novel normative modeling approach. Neuroimage Clin. 2021;30:102565.

11. Jain P, Ochi A, McInnis C, et al. Surgical outcomes in children with bottom-of-sulcus dysplasia and drug-resistant epilepsy: a retrospective cohort study. J Neurosurg Pediatr. 2021;28:1–11.

12. Macdonald-Laurs E, Warren AEL, Francis P, et al. The clinical, imaging, pathological and genetic landscape of bottom-of-sulcus dysplasia. Brain. 2023.

13. David B, Kröll-Seger J, Schuch F, et al. External validation of automated focal cortical dysplasia detection using morphometric analysis. Epilepsia. 2021;62:1005–1021.

14. Wagner J, Weber B, Urbach H, Elger CE, Huppertz H-J. Morphometric MRI analysis improves detection of focal cortical dysplasia type II. Brain. 2011;134:2844–2854.

15. Spitzer H, Ripart M, Whitaker K, et al. Interpretable surface-based detection of focal cortical dysplasias: a Multi-centre Epilepsy Lesion Detection study. Brain. 2022;145:3859–3871.

16. Adler S, Wagstyl K, Gunny R, et al. Novel surface features for automated detection of focal cortical dysplasias in paediatric epilepsy. NeuroImage: Clinical. 2017;14:18–27.

17. Gill RS, Lee H-M, Caldairou B, et al. Multicenter validation of a deep learning detection algorithm for focal cortical dysplasia. Neurology. 2021;97:e1571–e1582.

18. Zheng R, Chen R, Chen C, et al. Automated detection of focal cortical dysplasia based on magnetic resonance imaging and positron emission tomography. Seizure: European Journal of Epilepsy. 2024;117:126–132.

19. Harvey AS, Mandelstam SA, Maixner WJ, et al. The surgically remediable syndrome of epilepsy associated with bottom-of-sulcus dysplasia. Neurology. 2015;84:2021–2028.

20. Dale AM, Fischl B, Sereno MI. Cortical surface-based analysis. I. Segmentation and surface reconstruction. Neuroimage. 1999;9:179–194.

21. Berlangieri SU, Mito R, Semmelroch M, Pedersen M, Jackson G. Bottom-of-sulcus dysplasia: the role of 18F-FDG PET in identifying a focal surgically remedial epileptic lesion. European Journal of Hybrid Imaging. 2020;4:23.

22. Tournier JD, Smith R, Raffelt D, et al. MRtrix3: A fast, flexible and open software framework for medical image processing and visualisation. Neuroimage. 2019;202:116137.

23. Macdonald-Laurs E, Warren AEL, Lee WS, et al. Intrinsic and secondary epileptogenicity in focal cortical dysplasia type II. Epilepsia. 2023;64:348–363.

24. Greve DN, Van der Haegen L, Cai Q, et al. A surface-based analysis of language lateralization and cortical asymmetry. J Cogn Neurosci. 2013;25:1477–1492.

25. Spitzer H, Ripart M, Whitaker K, et al. Interpretable surface-based detection of focal cortical dysplasias: a Multi-centre Epilepsy Lesion Detection study. Brain. 2022;145:3859–3871.

26. Wang Y, Mo J, Sun Y, et al. Establishment of a normal control model of children’s brain 18-fluorodeoxyglucose positron emission tomography and analysis of the changing pattern in patients aged 0-14 years. Quant Imaging Med Surg. 2024;14:4703–4713.

27. Desikan RS, Ségonne F, Fischl B, et al. An automated labeling system for subdividing the human cerebral cortex on MRI scans into gyral based regions of interest. Neuroimage. 2006;31:968–980.

28. Wong T-T. Performance evaluation of classification algorithms by k-fold and leave-one-out cross validation. Pattern recognition. 2015;48:2839–2846.

29. Desarnaud S, Mellerio C, Semah F, et al. (18)F-FDG PET in drug-resistant epilepsy due to focal cortical dysplasia type 2: additional value of electroclinical data and coregistration with MRI. Eur J Nucl Med Mol Imaging. 2018;45:1449–1460.

30. Kikuchi K, Togao O, Yamashita K, et al. Diagnostic accuracy for the epileptogenic zone detection in focal epilepsy could be higher in FDG-PET/MRI than in FDG-PET/CT. Eur Radiol. 2021;31:2915–2922.

31. Tan Y-L, Kim H, Lee S, et al. Quantitative surface analysis of combined MRI and PET enhances detection of focal cortical dysplasias. NeuroImage. 2018;166:10–18.

32. Mo JJ, Zhang JG, Li WL, et al. Clinical Value of Machine Learning in the Automated Detection of Focal Cortical Dysplasia Using Quantitative Multimodal Surface-Based Features. Front Neurosci. 2018;12:1008.

33. Takaya S, Mikuni N, Mitsueda T, et al. Improved cerebral function in mesial temporal lobe epilepsy after subtemporal amygdalohippocampectomy. Brain. 2008;132:185–194.

34. Lagarde S, Boucekine M, McGonigal A, et al. Relationship between PET metabolism and SEEG epileptogenicity in focal lesional epilepsy. Eur J Nucl Med Mol Imaging. 2020;47:3130–3142.

35. Kudr M, Krsek P, Marusic P, et al. SISCOM and FDG-PET in patients with non-lesional extratemporal epilepsy: correlation with intracranial EEG, histology, and seizure outcome. Epileptic Disord. 2013;15:3–13.

36. Blumcke I, Cendes F, Miyata H, Thom M, Aronica E, Najm I. Toward a refined genotype-phenotype classification scheme for the international consensus classification of Focal Cortical Dysplasia. Brain Pathol. 2021;31:e12956.

37. Wang Y, Yu T, Blümcke I, et al. The clinico-pathological characterisation of focal cortical dysplasia type IIb genetically defined by MTOR mosaicism. Neuropathol Appl Neurobiol. 2023;49:e12874.

38. Zhao B, McGonigal A, Hu W, et al. Interictal HFO and FDG-PET correlation predicts surgical outcome following SEEG. Epilepsia. 2023;64:667–677.

39. Jin B, Lv Z, Chen W, et al. Perilesional white matter integrity in drug-resistant epilepsy related to focal cortical dysplasia. Seizure. 2021;91:484–489.

40. Labate A, Cherubini A, Tripepi G, et al. White matter abnormalities differentiate severe from benign temporal lobe epilepsy. Epilepsia. 2015;56:1109–1116.

41. Urquia-Osorio H, Pimentel-Silva LR, Rezende TJR, et al. Superficial and deep white matter diffusion abnormalities in focal epilepsies. Epilepsia. 2022;63:2312–2324.

42. Jin B, Krishnan B, Adler S, et al. Automated detection of focal cortical dysplasia type II with surface-based magnetic resonance imaging postprocessing and machine learning. Epilepsia. 2018;59:982–992.

43. Liang W, Tadesse GA, Ho D, et al. Advances, challenges and opportunities in creating data for trustworthy AI. Nature Machine Intelligence. 2022;4:669–677.

44. Wagstyl K, Adler S, Pimpel B, et al. Planning stereoelectroencephalography using automated lesion detection: Retrospective feasibility study. Epilepsia. 2020;61:1406–1416.

45. Fortin J-P, Cullen N, Sheline YI, et al. Harmonization of cortical thickness measurements across scanners and sites. NeuroImage. 2018;167:104–120.

