## Supplementary Table 2 for "Automated detection of bottom-of-sulcus dysplasia on MRI-PET in patients with drug-resistant focal epilepsy"

**Supplementary table 2: Calculated surface-based features**

| **Surface-based feature** | **Feature description** |
| --- | --- |
| *Cortical morphology* | |
| Mean curvature^46^ | Mean curvature was measured at the pial boundary as 1/r where r is the radius of an inscribed circle and is equal to the mean of the principals of curvature *k*_1_ and *k*_2_ (unit: 1/mm). An increase in mean curvature means the radius of curvature has decreased and folding is increased. This feature was not smoothed. |
| Local cortical deformation^47^ | Local cortical deformation was calculated as the product of the principal curvatures *k*_1_ and *k*_2_ and smoothed using a 10mm FWHM Gaussian kernel. |
| Sulcal depth | The integrated dot product of the movement vector of the cortical surface during inflation was used to calculate sulcal depth. The feature was not smoothed. |
| Local gyrification index^48^ | A measure of the amount of cortical surface invaginated in sulci within a circular region of interest smoothed using a 10mm FWHM Gaussian kernel |
| Cortical thickness^49^ | Cortical thickness was computed as the mean of the distance between the pial and white matter boundaries with an upper limit of 10mm and smoothed using a 10mm FWHM Gaussian kernel. |
| T1 grey-to-white matter  contrast^50^ | Grey matter was sampled at 30% cortical thickness (0.3) and white matter was sampled at a constant distance of 1mm and smoothed using a 10mm FWHM Gaussian kernel. |
| T1 signal  (cortical grey matter) | T1 signal was sampled between the pial and white matter boundaries at 20-80% of the cortical thickness in 10% increments to provide a mean T1 value and smoothed using a 10mm FWHM Gaussian kernel. |
| T1 signal  (subcortical grey matter) | Subcortical T1 signal was sampled 10mm below the grey-white matter boundary and smoothed using a 10mm FWHM Gaussian kernel |
| FLAIR signal  (cortical grey matter) | FLAIR signal was sampled between the pial and white matter boundaries at 20-80% of the cortical thickness in 10% increments to provide a mean FLAIR value and smoothed using a 10mm FWHM Gaussian kernel. |
| FLAIR signal  (subcortical white matter) | Subcortical FLAIR signal was sampled at 10mm below the white matter boundary and smoothed using a 10mm FWHM Gaussian kernel. |
| ^18^F-FDG-PET intensity (cortical grey matter)^51^ | ^18^F-FDG-PET signal was sampled between the pial and white matter boundaries at 20-80% of the cortical thickness in 10% increments to provide a mean ^18^F-FDG-PET value and smoothed using a 10mm FWHM Gaussian kernel. |
| ^18^F-FDG-PET intensity (subcortical grey matter) | ^18^F-FDG-PET was aligned to T1 and projected to the Freesurfer cortical surface. ^18^F-FDG-PET signal was sampled 10mm below the grey-white matter boundary and smoothed using a 10mm FWHM Gaussian kernel. |
