## Supplementary Table 1 for "Automated detection of bottom-of-sulcus dysplasia on MRI-PET in patients with drug-resistant focal epilepsy"

**Supplementary Table 1: Scanner and BOSD characteristics in training and test sets**

| **Patient number** | **Set** | **MRI scanner** | **PET scanner** | **MRI positive^a^** | **BOSD location** | **BOSD volume (cm^3^)** | **Pathologically proven** | **PET-MRI detector overlap with BOSD^b^**  **(top cluster)^c^** | **PET-only detector cluster overlap with BOSD (top cluster)** | **MRI-only cluster overlap with BOSD (top cluster)** |
| --- | --- | --- | --- | --- | --- | --- | --- | --- | --- | --- |
| 1 | Training | Siemens Trio Trim | Phillips Allegro | no | L post central sulcus (inf) | 4.69 | FCDIIB | yes  (yes) | yes  (yes) | yes  (yes) |
| 2 | Training | Siemens Trio Trim | Phillips Allegro | no | R lateral fissure/circular sulcus (post) | 1.39 | FCDIIB | yes  (yes) | yes  (yes) | yes  (yes) |
| 3 | Training | Siemens Trio Trim | Phillips Gemini TF 64 | no | L superior frontal gyrus (ant) | 3.0 | FCDIIB | yes  (yes) | yes  (yes) | yes  (yes) |
| 4 | Training | Siemens Trio Trim | Phillips Allegro | yes | R superior frontal gyrus (anteromedial) | 1.98 | FCDIIB | yes  (yes) | yes  (yes) | yes  (yes) |
| 5 | Training | Siemens Trio Trim | Phillips Allegro | yes | L cingulate sulcus (post) | 2.23 | FCDIIB | yes  (yes) | yes  (yes) | yes  (yes) |
| 6 | Training | Siemens Trio Trim | Phillips Allegro | no | L middle frontal gyrus (mid) | 0.39 | FCDIIB | no  (no) | no  (no) | no  (no) |
| 7 | Training | Siemens Trio Trim | Phillips Allegro | no | R middle frontal gyrus (anterior) | 1.27 | FCDIIB | yes  (yes) | yes  (yes) | yes  (yes) |
| 8 | Training | Siemens Trio Trim | Phillips Allegro | no | L cingulate sulcus (ant) | 1.76 | FCDIIA | yes  (yes) | yes  (yes) | no  (no) |
| 9 | Training | Siemens Trio Trim | Phillips Allegro | no | R cingulate sulcus (ant) | 3.6 | FCDIIB | yes  (yes) | yes  (yes) | no  (no) |
| 10 | Training | Siemens Verio | Phillips Ingenuity | no | L ascending ramus of lateral fissure | 0.79 | FCDIIA | yes  (yes) | yes  (yes) | yes  (yes) |
| 11 | Training | Siemens Trio Trim | Siemens Biograph128 | yes | R postcentral sulcus (inf) | 2.11 | FCDIIB | yes  (yes) | yes  (yes) | yes  (yes) |
| 12 | Training | Siemens Trio Trim | Siemens Biograph128 | no | L parieto-occipital sulcus | 9.11 | FCDIIB | yes  (yes) | yes  (yes) | yes  (yes) |
| 13 | Training | Siemens Trio Trim | Phillips Gemini | no | L circular sulcus (ant) | 1.04 | FCDIIB | yes (no) | yes  (yes) | no  (no) |
| 14 | Training | Siemens Biograph mMR | Siemens Biograph mMR | no | L superior parietal lobule | 2.30 | FCDIIA | yes  (yes) | yes  (yes) | yes  (yes) |
| 15 | Training | Siemens Biograph mMR | Siemens Biograph mMR | no | L inferior parietal lobule | 2.29 | FCDIIA | yes  (yes) | yes  (yes) | yes (no) |
| 16 | Training | Siemens Biograph mMR | Siemens Biograph mMR | no | R inferior frontal gyrus (ant) | 1.23 | FCDIIB | yes  (yes) | yes  (yes) | yes  (yes) |
| 17 | Training | Siemens Biograph mMR | Siemens Biograph mMR | no | R frontal pole (ant) | 1.06 | FCDIIA | yes (no) | yes (no) | yes  (yes) |
| 18 | Training | Siemens Biograph mMR | Siemens Biograph mMR | no | L middle frontal gyrus (post) | 0.09 | FCDIIB | yes (no) | yes  (yes) | no  (no) |
| 19 | Training | Siemens Trio Trim | Phillips Allegro | no | L precentral sulcus | 0.19 | FCDIIA | no (no) | no (no) | no  (no) |
| 20 | Training | Siemens Trio Trim | Phillips Allegro | no | L precentral sulcus | 0.48 | FCDIIA | yes (no) | yes  (yes) | no  (no) |
| 21 | Training | Siemens Biograph mMR | Siemens Biograph mMR | no | R middle temporal gyrus (post) | 0.49 | FCDIIB | yes  (yes) | yes  (yes) | no  (no) |
| 22 | Training | Siemens Biograph mMR | Siemens Biograph mMR | no | L pars triangularis | 0.57 | not operated | yes  (yes) | yes  (yes) | yes  (yes) |
| 23 | Training | Siemens Biograph mMR | Siemens Biograph mMR | no | R superior frontal gyrus (ant/pole) | 0.26 | not operated | yes (no) | yes  (yes) | no  (no) |
| 24 | Training | Siemens Biograph mMR | Siemens Biograph128 | yes | L superior temporal sulcus (post) | 2.76 | FCDIIB | yes  (yes) | yes  (yes) | yes  (yes) |
| 25 | Training | Siemens Biograph mMR | Siemens Biograph mMR | no | R precentral gyrus (inf) | 0.35 | FCDIIA | yes  (no) | yes  (yes) | no  (no) |
| 26 | Training | Siemens Biograph mMR | Siemens Biograph mMR | no | L precentral suclus (inf) | 0.93 | FCDIIB | yes  (yes) | yes  (yes) | yes  (yes) |
| 27 | Training | Siemens Biograph mMR | Siemens Biograph mMR | no | R precentral sulcus | 2.21 | FCDIIA | yes  (yes) | yes  (yes) | yes  (yes) |
| 28 | Training | Siemens Biograph mMR | Siemens Biograph mMR | no | R middle frontal gyrus | 2.53 | not operated | yes  (yes) | yes  (yes) | yes  (yes) |
| 29 | Training | Siemens Biograph mMR | Siemens Biograph mMR | no | L circular sulcus (post) | 4.11 | FCDIIA | yes  (yes) | yes  (yes) | yes  (yes) |
| 30 | Training | Siemens Biograph mMR | Siemens Biograph mMR | yes | L postcentral sulcus (inf) | 4.67 | FCDIIA | yes  (yes) | no (no) | yes  (yes) |
| 31 | Training | Siemens Biograph mMR | Siemens Biograph mMR | no | R circular sulcus (ant) | 1.75 | FCDIIA | yes  (yes) | yes  (yes) | yes  (yes) |
| 32 | Training | Siemens Biograph mMR | Siemens Biograph mMR | no | R superior temporal sulcus | 3.74 | FCDIIB | yes  (yes) | yes  (yes) | yes  (yes) |
| 33 | Training | Siemens Biograph mMR | Siemens Biograph mMR | no | R post central gyrus (sup) | 0.70 | FCDIIA | no (no) | no (no) | no  (no) |
| 34 | Training | Siemens Biograph | Siemens Biograph128 | no | L circular sulcus (mid) | 0.55 | FCDIIA | yes  (yes) | yes  (yes) | no  (no) |
| 35 | Training | Siemens Biograph mMR | Siemens Biograph mMR | no | L precentral gyrus | 0.11 | not operated | yes (no) | no (no) | no  (no) |
| 36 | Training | Siemens Biograph mMR | Siemens Biograph mMR | no | L precentral gyrus | 0.44 | not operated | yes  (yes) | no (no) | yes  (yes) |
| 37 | Training | Siemens Biograph mMR | Siemens Biograph mMR | no | R postcentral sulcus | 0.47 | not operated | yes  (yes) | yes  (yes) | yes  (yes) |
| 38 | Training | Siemens Biograph mMR | Siemens Biograph mMR | no | R superior temporal gyrus / supramarginal gyrus | 0.19 | FCDIIA | yes  (yes) | yes  (yes) | no  (no) |
| 39 | Training | Siemens Biograph mMR | Siemens Biograph mMR | no | L circular sulcus | 3.01 | not operated | yes  (no) | yes  (no) | no  (no) |
| 40 | Training | Siemens Biograph mMR | Siemens Biograph mMR | no | R cingulate sulcus (ant) | 0.50 | not operated | no  (no) | yes  (no) | no  (no) |
| 41 | Training | Siemens Biograph mMR | Siemens Biograph mMR | yes | R circular sulcus (ant) | 3.71 | not operated | yes  (yes) | yes  (yes) | yes  (yes) |
| 42 | Training | Siemens Biograph mMR | Siemens Biograph mMR | no | L postcentral sulcus | 0.48 | not operated | no  (no) | no  (no) | no  (no) |
| 43 | Training | Siemens Biograph mMR | Siemens Biograph mMR | yes | R middle frontal gyrus (post) | 2.65 | FCDIIB | yes  (yes) | yes  (no) | yes  (yes) |
| 44 | Training | Siemens Biograph mMR | Siemens Biograph mMR | no | L central sulcus (inf) | 1.97 | not operated | yes  (yes) | yes  (yes) | yes  (yes) |
| 45 | Training | Siemens Biograph mMR | Siemens Biograph mMR | no | L superior frontal sulcus (inf) | 0.58 | FCDIIB | yes  (no) | yes  (yes) | yes  (yes) |
| 46 | Training | Siemens Biograph mMR | Siemens Biograph mMR | no | R superior frontal gyrus (medial, post) | 0.27 | FCDIIB | yes  (yes) | yes  (yes) | yes  (yes) |
| 47 | Training | Siemens Biograph mMR | Siemens Biograph mMR | yes | R precentral sulcus (inf) | 5.77 | FCDIIB | yes  (yes) | yes  (yes) | yes  (yes) |
| 48 | Training | Siemens Trio Trim | Phillips Allegro | no | L supramarginal gyrus | 0.10 | operated, no pathology, SF>12m^e^ | yes  (yes) | yes  (yes) | no  (no) |
| 49 | Training | Siemens Biograph mMR | Siemens Biograph mMR | no | R posterior circular sulcus | 0.02 | FCDIIA | no  (no) | yes  (no) | no  (no) |
| 50 | Training | Siemens Biograph mMR | Siemens Biograph mMR | yes | R precentral sulcus (sup) | 0.32 | operated, no pathology, SF>12m | yes  (yes) | yes  (yes) | no  (no) |
| 51 | Training | Siemens Biograph mMR | Siemens Biograph mMR | no | L superior frontal sulcus | 0.81 | N/A | yes  (yes) | yes  (yes) | yes  (yes) |
| 52 | Training | Siemens Biograph mMR | Siemens Biograph mMR | no | R marginal sulcus | 1.36 | FCDIIA | yes  (yes) | yes  (yes) | yes  (yes) |
| 53 | Training | Siemens Biograph mMR | Siemens Biograph mMR | yes | R superior frontal gyrus (mid) | 2.21 | FCDIIA | no (no) | no (no) | no  (no) |
| 54 | Training | Siemens Biograph mMR | Siemens Biograph mMR | no | R parieto-occipital sulcus | 1.51 | not operated | yes (no) | no (no) | yes  (yes) |
| 55 | Test (RCH) | Siemens Biograph mMR | Siemens Biograph mMR | no | L inferior frontal gyrus | 0.55 | thermocoagulation, no pathology, SF>12m | yes  (yes) | yes  (yes) | yes  (yes) |
| 56 | Test (RCH) | Siemens Biograph mMR | Siemens Biograph mMR | no | L precentral gyrus | 0.19 | FCDIIA | yes  (yes) | yes  (yes) | no (no) |
| 57 | Test (RCH) | Siemens Biograph mMR | Siemens Biograph mMR | no | L precentral gyrus | 1.97 | not operated | yes  (yes) | no (no) | yes (no) |
| 58 | Test (RCH) | Siemens Biograph mMR | Siemens Biograph mMR | no | L anterior circular sulcus | 1.99 | not operated | yes  (yes) | yes  (no) | no (no) |
| 59 | Test (RCH) | Siemens Biograph mMR | Siemens Biograph mMR | no | R inferior frontal gyrus | 0.30 | FCDIIB | yes  (no) | yes  (yes) | no (no) |
| 60 | Test (RCH) | Siemens Biograph mMR | Siemens Biograph mMR | no | R precentral sulcus | 0.11 | not operated | no  (no) | no  (no) | no  (no) |
| 61 | Test (RCH) | Siemens Biograph mMR | Siemens Biograph mMR | no | R precuneus | 0.12 | FCDIIB | no  (no) | yes  (no) | no  (no) |
| 62 | Test (RCH) | Siemens Biograph mMR | Siemens Biograph mMR | no | R frontopolar | 2.35 | not operated | yes  (yes) | yes  (yes) | no  (no) |
| 63 | Test (RCH) | Siemens Biograph mMR | Siemens Biograph mMR | yes | R orbitofrontal | 0.64 | not operated | yes  (yes) | yes  (yes) | no  (no) |
| 64 | Test (RCH) | Siemens Biograph mMR | Siemens Biograph mMR | yes | L posterior circular sulcus | 4.78 | not operated | yes  (yes) | yes  (yes) | yes  (yes) |
| 65 | Test (RCH) | Siemens Biograph mMR | Siemens Biograph mMR | no | L superior frontal sulcus | 0.53 | FCDIIA | yes  (yes) | yes  (yes) | yes  (yes) |
| 66 | Test (RCH) | Siemens Biograph mMR | Siemens Biograph mMR | no | R anterior cingulate | 0.91 | LITT, no pathology, SF<12m^f^ | no  (no) | no  (no) | no  (no) |
| 67 | Test (RCH) | Siemens Biograph mMR | Siemens Biograph mMR | yes | R superior frontal sulcus | 0.82 | FCDIIA | yes  (yes) | yes  (yes) | yes  (no) |
| 68 | Test (RCH) | Siemens Biograph mMR | Siemens Biograph mMR | yes | R superior frontal sulcus | 0.28 | FCDIIA | yes  (yes) | yes  (yes) | no  (no) |
| 69 | Test (RCH) | Siemens Biograph mMR | Siemens Biograph mMR | no | R frontopolar | 0.92 | not operated | yes  (yes) | yes  (yes) | no  (no) |
| 70 | Test (RCH) | Siemens Biograph mMR | Siemens Biograph mMR | no | R anterior circular sulcus | 1.58 | not operated | yes  (yes) | yes  (yes) | yes  (yes) |
| 80 | Test (RCH) | Siemens Biograph mMR | Siemens Biograph mMR | no | R medial superior frontal gyrus | 0.43 | not operated | no  (no) | no  (no) | no  (no) |
| 81 | Test  (Austin) | Siemens Magnetom Skyra | Phillips Allegro | unknown | R precentral sulcus | 1.03 | FCDIIA | no  (no) | no  (no) | no  (no) |
| 82 | Test  (Austin) | Siemens Trio Trim | Phillips Allegro | unknown | R superior parietal lobule | 3.66 | FCDIIB | no  (yes) | yes  (yes) | no  (no) |
| 83 | Test  (Austin) | Siemens Trio Trim | Phillips Allegro | unknown | L middle frontal gyrus | 3.17 | FCDIIB | yes  (yes) | yes  (yes) | yes  (yes) |
| 84 | Test  (Austin) | Siemens Magnetom Avanto | Phillips Allegro | unknown | R frontopolar | 4.44 | FCDIIB | yes  (yes) | yes  (yes) | yes  (yes) |
| 85 | Test  (Austin) | Siemens Magnetom Avanto | Phillips Allegro | unknown | R frontopolar | 1.32 | FCDIIB | yes  (yes) | yes  (yes) | yes  (yes) |
| 86 | Test  (Austin) | Siemens Magnetom Skyra | Phillips Allegro | unknown | R middle frontal gyrus | 8.18 | FCDIIB | yes  (yes) | yes  (yes) | yes  (yes) |
| 87 | Test  (Austin) | Siemens Trio Trim | Phillips Allegro | unknown | L anterior cingulate | 0.45 | FCDIIB | no  (no) | no  (no) | no  (no) |
| 88 | Test  (Austin) | Siemens Magnetom Syra | Phillips Gemini TF 64 | unknown | L superior frontal gyrus | 1.92 | FCDIIB | yes  (no) | yes  (no) | no  (no) |
| 89 | Test  (Austin) | Siemens Magnetom Skyra | Phillips Gemini TF 64 | unknown | R precuneus | 1.75 | FCDIIB | yes  (no) | yes  (no) | no  (no) |
| 90 | Test  (Austin) | Siemens Magnetom Avanto | Phillips Allegro | unknown | R postcentral sulcus | 6.17 | FCDIIB | yes  (no) | yes  (no) | no  (no) |
| 91 | Test  (Austin) | Siemens Magnetom Skyra | Phillips Gemini TF 64 | unknown | L frontal pole | 0.44 | FCDIIB | yes  (no) | no  (no) | yes  (yes) |
| 92 | Test  (Austin) | Siemens Magnetom Avanto | Phillips Gemini TF 64 | unknown | L orbitofrontal cortex | 1.56 | FCDIIB | yes  (no) | yes  (no) | yes  (no) |

ant = anterior, Austin = Austin Hospital, FCDIIA = Focal cortical dysplasia type IIA, FCDIIB = focal cortical dysplasia type IIB, inf = inferior, L = left, LITT = laser interstitital thermal therapy, post = posterior, R= right, RCH = Royal Children’s Hospital, sup = superior, SF>12m = seizure free more than 12 months, SF<12m = seizure free less than 12 months

^a^MRI positive based on initial scan which may have been performed outside of the Royal Children’s or Austin Hospital,  ^b^overlap with any of the top five clusters on the MRI-PET BOSD detector, ^c^overlap with the top cluster on the MRI-PET BOSD detector
